# Application of Urinary Peptide-Biomarkers in Trauma Patients as a Predictive Tool for Prognostic Assessment, Treatment Interventions, and Intervention Timing: Prospective Nonrandomized Pilot Study

**DOI:** 10.1101/2024.07.24.24310868

**Authors:** Gökmen Aktas, Felix Keller, Justyna Siwy, Agnieszka Latosinska, Harald Mischak, Jorge Mayor, Jan Clausen, Michaela Wilhelmi, Vesta Brauckmann, Stephan Sehmisch, Tarek Omar Pacha

**Author notes:** Contributing authors. These authors contributed equally to this work.

## Abstract

**Background:** Treatment of severely injured patients represents a major challenge, in part due to the unpredictable risk of major adverse events, including death. Preemptive personalized treatment aimed at preventing these events is a key objective of patient management; however, the currently available scoring systems provide only moderate guidance. Molecular biomarkers from proteomics/peptidomics studies hold promise for improving the current situation, ultimately enabling precision medicine based on individual molecular profiles.

**Methods:** To test the hypothesis that proteomics biomarkers could predict patient outcomes in severely injured patients, we initiated a pilot study involving consecutive urine sampling (on days 0, 2, 5, 10, and 14) and subsequent peptidome analysis using capillary electrophoresis coupled to mass spectrometry (CE-MS) of 14 severely injured patients and two additional ICU patients. The urine peptidomes of these patients were compared to the urine peptidomes of age- and sex-matched controls. Previously established urinary peptide-based classifiers, CKD274, AKI204, and CoV50, were applied to the obtained peptidome data, and the association of the scores with a combined endpoint (death and/or kidney failure and/or respiratory insufficiency) was investigated.

**Results:** CE-MS peptidome analysis identified 281 peptides that were significantly altered in severely injured patients. Consistent upregulation was observed for peptides from A1AT, FETUA, and MYG, while peptides derived from CD99, PIGR and UROM were consistently reduced. Most of the significant peptides were from different collagens, and the majority were reduced in abundance. Two of the predefined peptidomic classifiers, CKD273 and AKI204, showed significant associations with the combined endpoint, which was not observed for the routine scores generally applied in the clinics.

**Conclusions:** This prospective pilot study confirmed the hypothesis that urinary peptides provide information on patient outcomes and may guide personalized interventions based on individual molecular changes. The results obtained allow the planning of a well-powered prospective trial investigating the value of urinary peptides in this context in more detail.

## Background

Severe trauma patients represent a significant medical challenge, often requiring intensive care unit (ICU) admission due to the life-threatening nature of their injuries (1). These patients have sustained critical injuries, e.g., car accidents, high-energy trauma or falls from great heights(2). Managing severely injured patients in the ICU involves a multidisciplinary approach(3). The outcome of these patients can be unpredictable given the severity of their injuries(4). Often, despite intensive monitoring and intervention, their condition can deteriorate significantly, become unsustainable, and ultimately result in death due to multiple organ dysfunction syndrome (MODS)(5).

When referring to polytraumatized patients, the definition by Muhr and Tscherne is frequently applied (6). However, the term ’polytrauma’ is used inconsistently. Therefore, a new definition of polytrauma was recently developed(7). Today, the widely accepted definition, the New Berlin Score, characterizes polytrauma as a significant injury in at least 2 body regions with an abbreviated injury scale (AIS) score of 3 or higher and at least one pathological value for one of the following parameters: age (above 70), hypotension (systolic blood pressure <90 mmHg), unconsciousness (Glasgow coma scale (GCS) at the scene <=8), acidosis (base excess <=-6), and coagulopathy (partial thromboplastin time (PTT) >=40/INR >=1.4)(7).

Similarly, the definition of ’multiorgan failure’ (MOF) has evolved. MOF was redefined in 1992 through a consensus conference(8). MOF was no longer defined as a static condition but rather a dynamic and reversible process. Additionally, the classical MOF was replaced with MODS and comprehensively defined. In this definition, MODS is described as the dysfunction of organs in critically ill patients, which can only be resolved through therapeutic interventions(8).

Mortality rates for MODS have improved over time in the US, decreasing from 33-36% in approximately 2010 to 22% in more recent studies(9,10). Of these deaths, 24% are due to cardiovascular events within the first few days of ICU admission(10).

Various scoring systems, including the sequential organ failure assessment (SOFA) score and the multiorgan dysfunction (MOD) score, which are common metrics in clinical studies of intensive care medicine, have been developed to assess the severity of organ dysfunction and predict mortality(11). Both systems meet the requirements for describing organ dysfunction. Both SOFA and MOD assess the same six organ systems, but they have practical differences in their calculations(11). The SOFA score is based on the most deviant value within a 24-hour period, while the MOD score uses physiological values measured at the same time every day to avoid capturing momentary physiological changes(11). The acute physiology and chronic health evaluation II (APACHE II) score provides further metrics and is commonly used complimentary to the SOFA score (1). In addition to parameters from individual organ systems, it incorporates the patient’s age, current clinical findings, and medical history to make predictions about the likelihood of survival in an intensive care setting(12).

The simplified acute physiology score II (SAPS II) allows for assessing the severity of disease in a patient admitted to an ICU and provides information about morbidity(13). Additionally, the score, in combination with the therapeutic intervention scoring system (TISS), can be used to assess the progression of disease severity and the intensity of treatment effort(13).

These scores help clinicians evaluate the severity of disease and the intensity of treatment needed, although they do not always account for trauma as the underlying cause of ICU admission.

Specifically, the injury severity score (ISS) and the trauma and injury severity score (TRISS) are widely used. The ISS is calculated by adding the three highest body area values of the abbreviated injury scale (AIS) squared together(14). The TRISS, introduced in 1981, is a combined index based on the revised trauma score (RTS), ISS, and patient age to predict survival probabilities(14). The results indicate that the TRISS aligns well with expected outcomes and performs favourably compared to other indices, such as the ISS(14).

However, these variables may not capture the full complexity of a patient’s condition.

The revised injury severity classification (RISC) score, introduced in 2003, included not only anatomical injury descriptions but also physiological indicators (e.g., base deficit, haemoglobin, and cardiopulmonary resuscitation)(15). To address some remaining issues, accompanied by the RISC score, the RISC II was developed(15). In addition to other changes, it includes new predictors such as pupil size and reactivity . The RISC-II model outperforms the original RISC, particularly in terms of discrimination, precision, and calibration. It can be used to estimate the risk of death in severely injured patients more effectively. The integration of RISC II into daily intensive care practice, especially as a tool for monitoring patient progress, is challenging due to its complex calculation and the collection of numerous parameters(15,16).

In the context of ICU management, advanced laboratory testing and monitoring are crucial. Parameters such as N-terminal pro B-Typ natriuretic peptide (NT-proBNP) and troponin are used to assess cardiac function(17). However, there is a known mutual influence of these parameters in cases of concurrent kidney failure, prompting more recent literature to emphasize more recent, independent markers such as B-type natriuretic peptide.

The emergence of "-omics" approaches, including proteomics and genomics, has potential for improved prediction and treatment strategies in trauma care. Changes in urine peptide patterns can provide valid insights into several organ systems (18,19). Biomarkers such as AKI204, which is useful for predicting acute kidney injury (AKI) in ICU patients (20), and CKD273, which is a biomarker for the detection of chronic kidney disease (CKD), can predict worsening of kidney function (21) or cardiovascular outcome (22) and may be good biomarkers for both predicting kidney damage in polytraumatized patients and providing the time to start renal replacement therapy early (23,24). The COV50 classifier, developed during the COVID-19 pandemic was able to predict the incidence of death and disease progression in infected patients (25). It also demonstrated significant prediction of mortality in patients without SARS-CoV-2 infection in the ICU but also in the general population (25,26). These results support the hypothesis that urinary peptide-based biomarker classifiers represent a significant advancement in the ability to monitor and manage complex pathophysiological processes in polytraumatized patients. Therefore, the aims of this pilot study were to examine whether trauma-specific peptides can be identified in urine samples and whether urinary peptide-based classifiers may predict patient outcomes and organ failure.

## Methods

### Cohort description

In this prospective nonrandomized pilot study, urinary proteomic data from 16 severely injured patients recruited between January and July 2023 were examined. All adult patients aged ≥18 years of both genders who had experienced primary trauma and were admitted to our level 1 trauma center were included. Patients with direct traumatic consequences to the urinary tract, bladder, or kidneys were excluded due to unpredictable analysis failure.

Treatment followed the current state of medical practice, in accordance with guidelines and international standards.

Each patient was informed about the study and the procedure either immediately or after regaining consciousness and capacity. In the case of a caregiver situation, the caregiver was provided with appropriate information. Informed consent was obtained retrospectively once a caregiver was available or the capacity for consent was established. If retrospective inclusion in the study was undesirable, the patient’s collected data were excluded. All participants provided written consent, and the study was approved by the Ethics Committee of Hannover Medical School (No: 10415_BO_S_2022).

The study was conducted in accordance with Good Clinical Practice and the Declaration of Helsinki. According to the New Berlin score, 14 of the 16 patients met the criteria for polytrauma (19). These multiple-injury patients had an ISS ≥ 16, at least two injuries with an AIS of ≥ 3 and an indication for intensive care monitoring or surgery. The other two patients had a single injury requiring a longer hospital stay (e.g. vacuum sealing, external fixator, extensive soft tissue damage) ensuring that the entire monitoring period was completed.

### Urine sample and data collection

For all 16 patients, urine samples and data were collected in the department of Trauma Surgery, Medical School of Hanover, Hanover, Germany. If a urinary catheter was inserted in the emergency room, the first urine sample was collected, as shown in the flow diagram (**Figure 1**). Blood was also sampled according to polytrauma standards. If patients were directly admitted to the intensive care unit following admission to the emergency room, the first urine sample was collected. According to the study design, urine was sampled on days 0, 2, 5, 10, and 14 using a urine monovette containing boric acid (Sarstedt, Nümbrecht, Germany) and stored frozen at -20°C.

**Figure 1:**
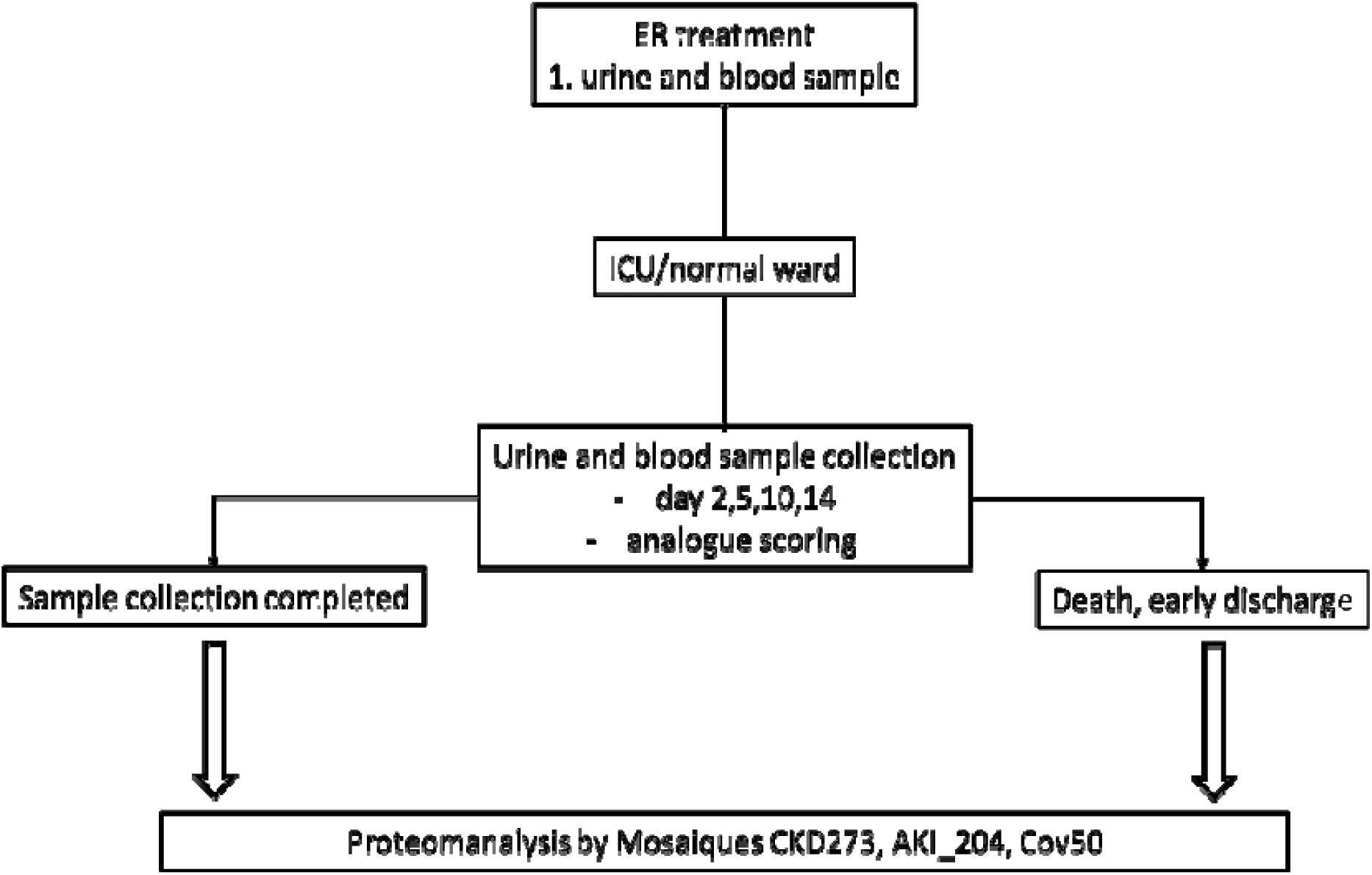
Collection of urine and blood samples and scoring

### Parameters

On the days of urine collection, the patients were assessed using current intensive care scores. On the day of admission, the ISS, TRISS, RISC II, APACHE-II, SAPS-II, and TIPS scores were recorded. Since the extent of injury did not change, the ISS and TRISS were only recorded at admission. APACHE-II was scored analogously on all sampling days, while SAPS-II and TIPS were assessed once upon admission.

Blood samples were also taken on the sampling days. The samples were investigated for electrolytes, lactate, CRP, complete blood count with haemoglobin and leukocytes, coagulation parameters (INR, PTT), and kidney retention parameters.

Furthermore, the following events were recorded: Long-term ventilation, tracheotomy, prone position, acute respiratory distress syndrome (ARDS), acute kidney failure, dialysis, cardiovascular events, resuscitation, death, emergency surgery, extracorporeal membrane oxygenation (ECMO), and early discharge from the ICU (**Supplementary Table 1**). The combined endpoint was defined as death and/or kidney failure and/or respiratory insufficiency.

### Proteomic analysis and data processing

Urine sample preparation was performed as previously described (27). Briefly, the urine samples were thawed immediately before use, and 0.1 ml of urine was diluted with 0.7 ml of 2 M urea and 10 mM NH4OH containing 0.02% SDS. The samples were ultrafiltered and desalted using a PD-10 column (GE Healthcare, Danderyd, Sweden) equilibrated with 0.01% NH4OH. The filtrate was lyophilized and stored at 4°C until Capillary electrophoresis–mass spectrometry (CE-MS) analysis was performed. CE-MS analyses were performed on a P/ACE MDQ capillary electrophoresis system (Beckman Coulter, Brea, California) coupled on-line to a micrOTOF II mass spectrometer (Bruker Daltonic, Bremen, Germany) as previously described (28).

The mass spectral peaks were deconvoluted using MosaFinder software (29). Normalization was performed using a linear regression algorithm with internal standard peptides as references. The detected peptides were deposited, matched, and annotated in the Microsoft SQL database (Microsoft, Redmond, Washington,USA).

The amino acid sequences were obtained by performing MS/MS analysis using a P/ACE CE coupled to a Q Exactive™ Plus Hybrid Quadrupole-Orbitrap™ MS instrument (Thermo Fisher Scientific, Waltham, Massachusetts, USA). The mass spectrometer was operated in data-dependent mode to automatically switch between MS and MS/MS acquisition. The data files were searched against the UniProt human nonredundant database using Proteome Discoverer 2.4 and the SEQUEST search engine without enzyme specification (activation type: HCD; precursor mass tolerance: 5 ppm) ; fragment mass tolerance: 0.05 Da). The false discovery rate (FDR) was set to 1%. For further validation of the obtained peptide sequences, the correlation between peptide charge at a working pH of 2 and CE migration time was used to minimize false-positive derivation rates (30). The calculated CE migration times of the sequence candidates based on their peptide sequences (number of basic amino acids) were compared to the experimental migration times.

### Application of urine peptide-based biomarker classifiers

In this study, predefined urinary peptide-based classifiers for predicting the onset and progression of CKD, AKI, and ARDS were used. For testing the endpoints, the classifiers CKD273, AKI204, and Cov50 were applied. previously(19,20,31).

### Statistics

Statistical analysis was performed using SPSS computer software (SPSS 28, IBM, Armonk, New York, USA). Demographics, clinical variables, and proteomics classification scores are summarized as the means±SDs, and categorical variables are presented as frequencies (%). Group means were compared using the Wilcoxon rank sum exact test, Fisher’s exact test and Wilcoxon rank sum test.

The CE-MS-based data of 14 trauma patients and 14 sex- and age-matched healthy controls extracted from the human urinary databasewere used to identify urinary peptides potentially associated with trauma (29). The statistical analysis was performed using R-based statistical software. Only peptides with a frequency threshold of at least 70% in one of the groups were considered. P values were obtained using the Wilcoxon signed-rank test, followed by Benjamini–Hochberg false discovery rate adjustment.

## Results

### Cohort characteristics

The patient characteristics of the whole cohort (n=16) are shown in **Table 1**. The average age of the study population was 46 ± 21 years. The mean ISS was 27 ± 15, the average TRISS was 56 ± 39, and the average RISC II score was 11±14. In **Table 2** mean AIS, systolic blood pressure and ASA score (classification of patients regarding their physical condition, American Society of Anesthesiologists) of the 14 polytraumatized patients are shown. The mean AIS for the head, face, thorax, abdomen, extremities with pelvic injuries and external injuries were 2.00±1.24, 0.64±0.84, 2.64±1.65, 2.00±1.74, 2.50±1.40 and 0.93±0,83, respectively, at admission. The mean systolic blood pressure was 133.93 mmHg±17.67 at admission. The average preclinical ASA score was 1.43. Upon admission, the average SAPS II score was 32 (±13), and the average TIPS score was 17.54 (±9.46).

**Table 1:**
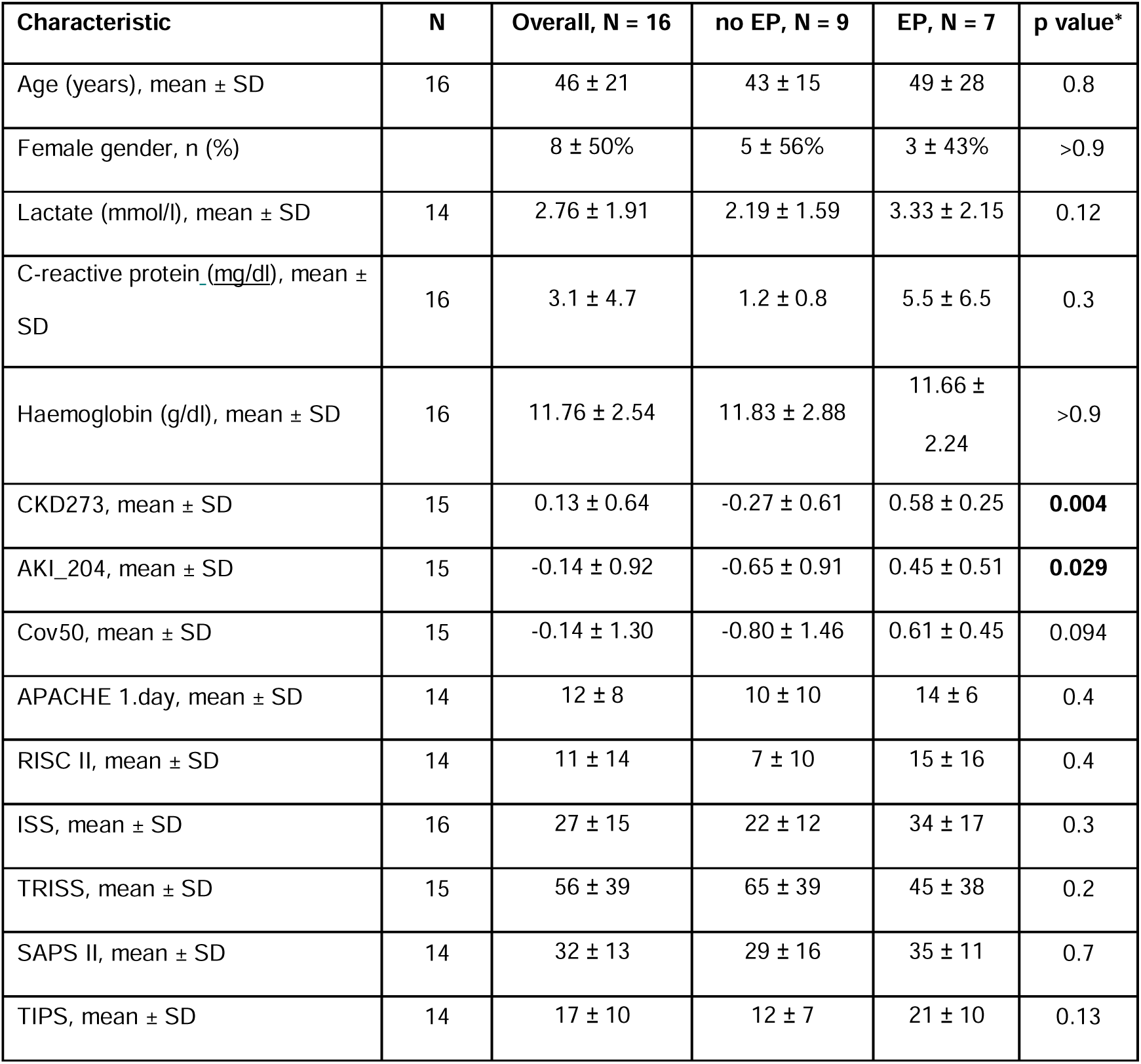
Mean values for baseline cohort characteristics of all 16 patients included (14 polytrauma patients), EP EP = Endpoint, *Wilcoxon rank sum exact test; Fisher’s exact test; Wilcoxon rank sum test.

| Characteristic | N | Overall, N = 16 | no EP, N = 9 | EP, N = 7 | p value* |
| --- | --- | --- | --- | --- | --- |
| Age (years), mean ± SD | 16 | $46 \pm 21$ | $43 \pm 15$ | $49 \pm 28$ | 0.8 |
| Female gender, n (%) | | $8 \pm 50\%$ | $5 \pm 56\%$ | $3 \pm 43\%$ | >0.9 |
| Lactate (mmol/l), mean ± SD | 14 | $2.76 \pm 1.91$ | $2.19 \pm 1.59$ | $3.33 \pm 2.15$ | 0.12 |
| C-reactive protein (mg/dl), mean ± SD | 16 | $3.1 \pm 4.7$ | $1.2 \pm 0.8$ | $5.5 \pm 6.5$ | 0.3 |
| Haemoglobin (g/dl), mean ± SD | 16 | $11.76 \pm 2.54$ | $11.83 \pm 2.88$ | $11.66 \pm 2.24$ | >0.9 |
| CKD273, mean ± SD | 15 | $0.13 \pm 0.64$ | $-0.27 \pm 0.61$ | $0.58 \pm 0.25$ | **0.004** |
| AKI_204, mean ± SD | 15 | $-0.14 \pm 0.92$ | $-0.65 \pm 0.91$ | $0.45 \pm 0.51$ | **0.029** |
| Cov50, mean ± SD | 15 | $-0.14 \pm 1.30$ | $-0.80 \pm 1.46$ | $0.61 \pm 0.45$ | 0.094 |
| APACHE 1.day, mean ± SD | 14 | $12 \pm 8$ | $10 \pm 10$ | $14 \pm 6$ | 0.4 |
| RISC II, mean ± SD | 14 | $11 \pm 14$ | $7 \pm 10$ | $15 \pm 16$ | 0.4 |
| ISS, mean ± SD | 16 | $27 \pm 15$ | $22 \pm 12$ | $34 \pm 17$ | 0.3 |
| TRISS, mean ± SD | 15 | $56 \pm 39$ | $65 \pm 39$ | $45 \pm 38$ | 0.2 |
| SAPS II, mean ± SD | 14 | $32 \pm 13$ | $29 \pm 16$ | $35 \pm 11$ | 0.7 |
| TIPS, mean ± SD | 14 | $17 \pm 10$ | $12 \pm 7$ | $21 \pm 10$ | 0.13 |

**Table 2:** Mean values for AIS, systolic bloodpressure and ASA of all 14 polytraumatzed patients.

| Characteristics | N | mean | SD |
| --- | --- | --- | --- |
| Abbreviated Injury Scale (AIS) | 14 | 2,00 | 1,24 |
| Head | 14 | 0,64 | 0,84 |
| Face | 14 | 2,64 | 1,65 |
| Thorax | 14 | 2,00 | 1,74 |
| Abdomen | 14 | 2,50 | 1,40 |
| Extremities, pelvis | 14 | 0,93 | 0,83 |
| External |  |  |  |
| Systolic bloodpressure on admission (mmHg) | 14 | 133,93 | 17,67 |
| ASA | 14 | 1,43 | 0,65 |

### Trauma patient outcomes

**Supplementary Table 1** lists the frequencies of injuries and recorded events during the 14- day monitoring period in the polytrauma patients (n=14). Eleven patients suffered from traumatic brain injury (n=11), while one patients had spinal cord injury. Thoracic trauma was present in 71% of the patients (n=10), and half of the patients had both abdominal trauma (n=7) and pelvic injuries (n=7). The following events were observed during the study: 1 patient did not survive the study (7%). Four patients developed respiratory insufficiency (29%), and an equal number of patients suffered acute kidney failure (29%), with 1 patient requiring dialysis. ECMO support was necessary for one patient (7%). Additionally, one patient needed to be placed in the prone position during intensive care (7%). ARDS developed in 1 patient (7%). **Supplementary Table 2** shows the frequencies of successful probe collection, score and bloodsample trends over the whole monitoring period of 14 days. During the monitoring period, not all urine samples could be collected due to early discharge. Eight patients completed the entire study in 14 days (57%). Despite early discharge and the reduction in the number of patients from the third sampling point (Day 5), the average APACHE II score showed no consistent trend over time (12.3 ±7.6, 11.2 ±6.0, 10.9 ±6.2, 12.1 ±6.2, 12.6 ±6,6). In the laboratory results corresponding to the sampling days, the highest average lactate levels were observed on the day of admission (2.8 mmol/l ±1.9).

Subsequently, there was a successive decrease, with normalization of the lactate levels on the following sampling days (1.6 mmol/l ±1.6, 1.0 mmol/l ±0.4, 0.8 mmol/l ±0.3, 0.7 mmol/l ±0.1).

On average, all included patients had normal CRP levels at admission (3.2 mg/dl ±1.9). At sampling days 2 and 4, increases in the average values of 1695 mg/dl ±99.46 and 139.0 mg/dl ±166.64, respectively, were observed.

On the day of admission, the patients had an average Hb level of 11.7 g/dl ±2.7, which showed the greatest decrease at sampling day 2 (9.1 g/dl ±2.2) and then a successive mild decline at the remaining sampling days (9.0 g/dl ±1.7, 8.3 g/dl ±1.3, 8.9 g/dl ±0.9). During the monitoring period, 3 patients did not require any packed red blood cells (PRBC) (21%), 1 patient received 4 PRBCs (7%), 4 patients each received 5 (29%), 2 patients required 7 (14%), another 2 patients received 12 (14%), and one patient required 36 while another needed 57 PRBCs (7%).

### Definition of urinary peptides specific for polytrauma patients

For the definition of urinary peptides significantly associated with trauma, the urinary peptide data of 14 polytraumatized patients and 14 age- and sex-matched healthy controls from the Human Urinary Peptide Database were used(32). This comparison revealed that 281 peptides were significantly affected (p<0.05, Wilcoxon test followed by adjustment for multiple comparisons by Benjamini and Hochberg). All the identified peptides are listed in **Supplementary Table 3**. We observed consistent upregulation of the expression of the peptides Alpha-1-antitrypsin (A1AT) (n=3), Fetuin (FETUA) (n=3), and Myoglobin (MYG) (n=2), which is in line with expectations and downregulation of the expression of fragments of CD99 (n=9), PIGR (n=9) and UROM (n=6). One highly upregulated ProSAAS fragment was also detected. Most of the significant peptides were fragments of different collagens (n=205), 118 of which were downregulated.

### Application of urinary peptide-based classifiers and their association with established severity scores

On the obtained CE-MS data, the previously developed classifiers CKD273, AKI204 and CoV50 were applied. For each classifier and patient, the numeric classification score was calculated.

The classifier for CKD prediction, CKD273, demonstrated a significant correlation with the ISS (p=0.034) and the APACHE-II score (p<0.001). The APACHE-II score was the only severity score that was obtained not only at baseline but also at every follow-up timepoint. The AKI204 classifier correlated significantly (p=0.031) with the SAPS and APACHE-II score (p<0.001). The Cov50 classifier was significantly correlated with the ISS (p=0.042), the RISC II (p=0.040) and the APACHE-II score (p<0.001). A chart of the correlation statistics is shown in **Figure 2**.

**Figure 2:**
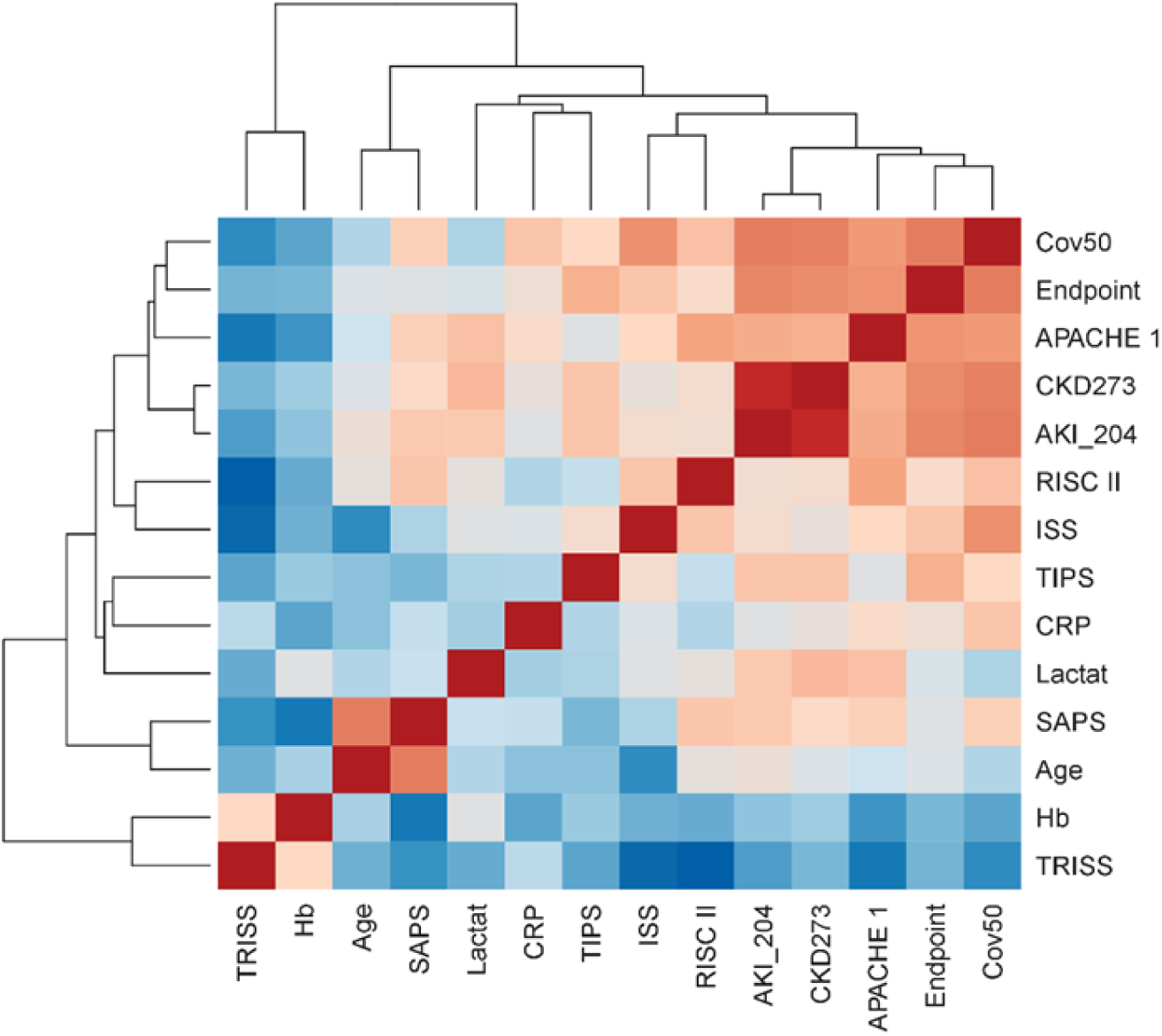
Chart of correlation statistics between the clinical parameters, severity scores and peptide-based classifiers.

### Associations of the parameters with the combined endpoint

In the next step, the associations of clinical parameters, severity scores, and the obtained peptide-based classification scores at baseline with the combined endpoint were investigated. The results are listed in **Table 1** and shown in the form of box-whisker plots in **Figure 3**. Significant differences between the patients who reached the combined endpoint and those who did not were observed for CKD273 and AKI204 (p= 0.004 and p= 0.029), where the Cov50 classifier showed only a trend (p=0.094). The clinical parameters and severity scores did not significantly differ between the groups. **Figure 4** displays the classification scores of the peptide-based classifiers throughout the entire study period.

**Figure 3:**
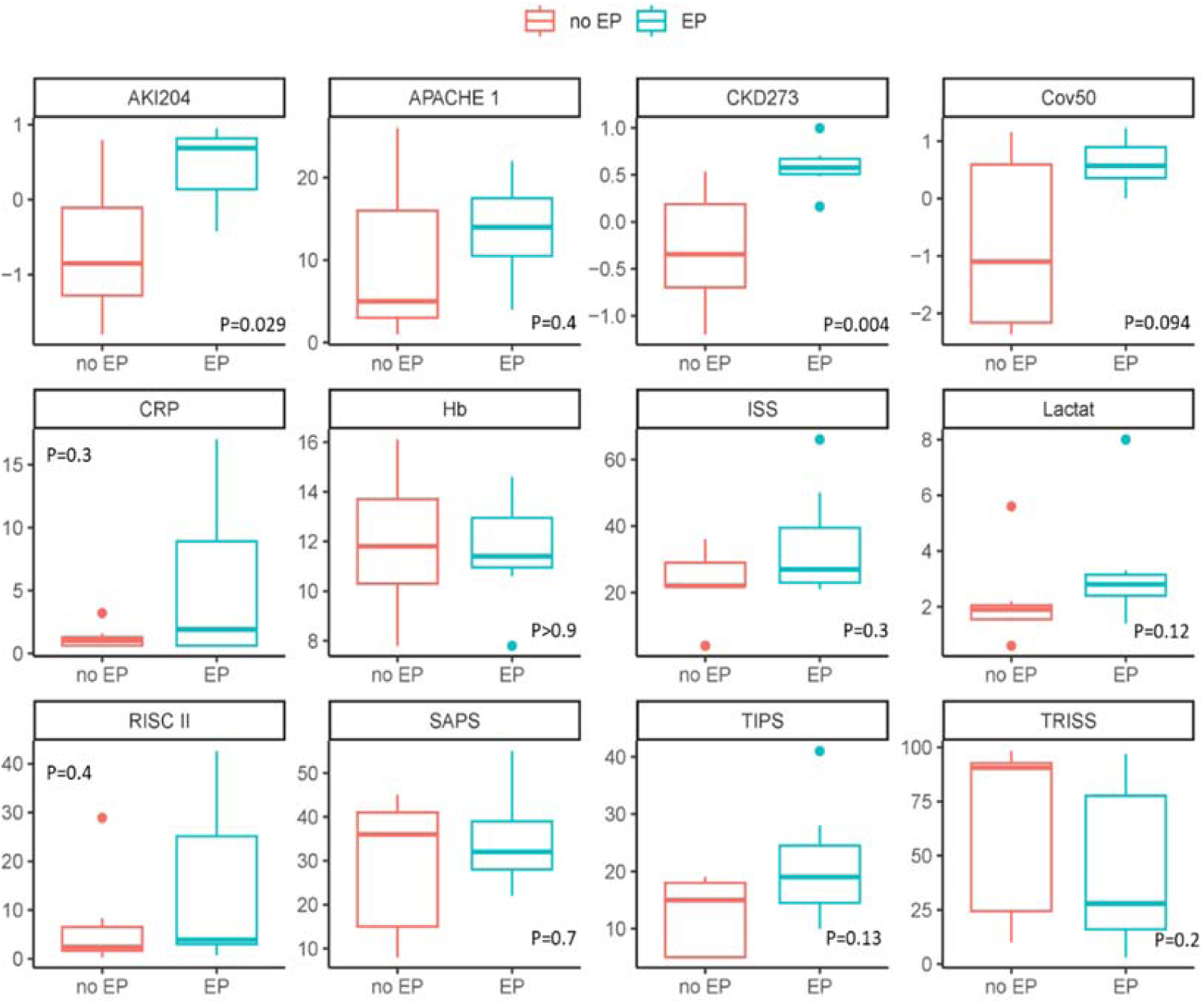
Association of the parameters with the combined endpoint. Box-whisker plots of the baseline parameters and their associations with the prediction of the combined endpoint (EP) are shown.

**Figure 4:**
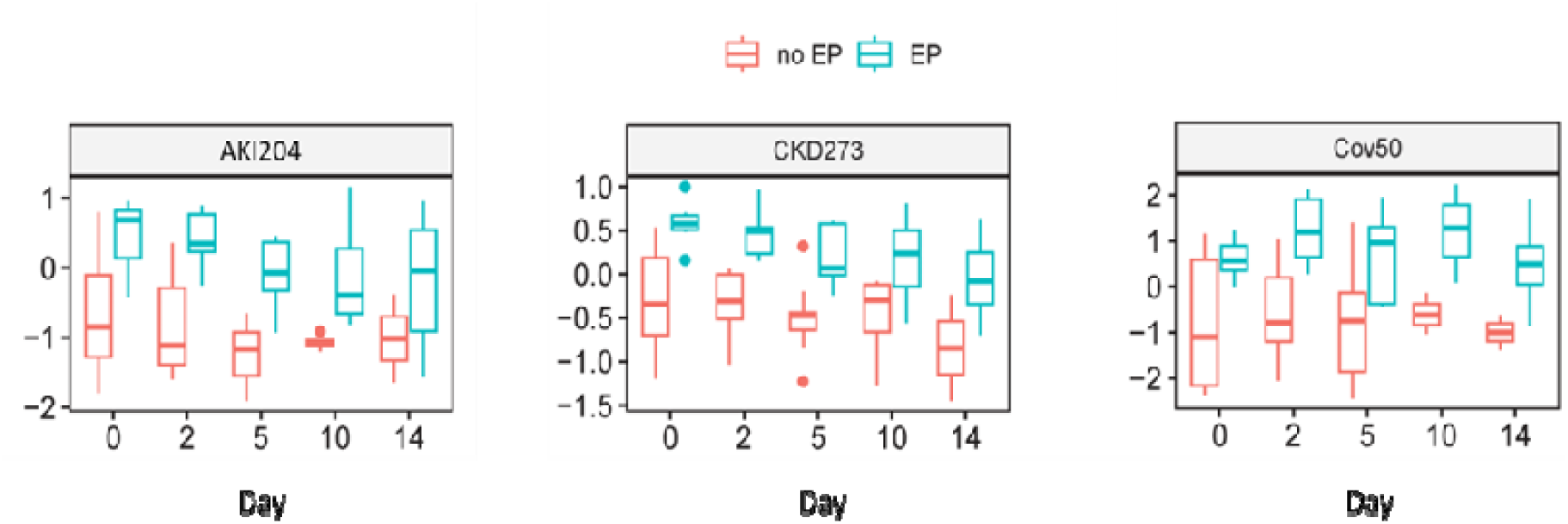
Distribution of classification scores obtained using the urinary peptide-based classifiers AKI204, CKD273, and Cov50 between patients who reached the combined endpoint and those who did not throughout the entire study duration.

## Discussion

The aim of this study was to investigate whether

a. severely injured trauma patients exhibit significant deregulation of urinary peptides, and
b. such changes can be used to predict outcomes and guide treatment.

The combined endpoints of the patients in this study were defined as death and/or kidney failure and/or respiratory insufficiency.

As hypothesized, our data demonstrated a highly significant deregulation of urinary proteom levels in trauma patients. A significant change was observed in CKD273 (p = 0.004) and in AKI_204 (p = 0.029). Cov50 showed a weak trend (p = 0.094).

These urine proteomes were initially used for predicting acute or acute-on-chronic kidney failure (AKI_207, CKD237) and the development of adverse outcomes in COVID-19 patients (Cov50) (20–22,33–35). Only one patient (7,1%) developed ARDS in this small cohort, which could explain the non-significant trend in Cov50 (p = 0.094). In contrast, four patients experienced acute kidney failure, and one patient required dialysis

Overall, 281 deregulated peptides were observed among the polytrauma patients. Among the measured peptides, the most notable changes were observed in collagen peptides (**Supplementary Table 3**). This was expected, as collagen peptides are the most abundant peptides in urine and are influenced by several pathologies, including kidney and cardiovascular diseases, as well as death (30). As shown here, the effect on collagen peptides was inconsistent across these pathologies; some peptides were significantly upregulated, while others were highly significantly and consistently reduced(**Supplementary Table 3**). Although these peptides are very common, the molecular mechanisms underlying their regulation and their connection to (patho)physiology remain unknown.

### Kidney failure

Alpha-1-antitrypsin and Fetuin showed significant deregulations (**Supplementary Table 3**). Similar changes have been observed previously in patients with chronic or acute kidney diseases, likely indicating severe renal stress in trauma patients (22,28,33). Stress or damage to the kidney is further indicated by the consistent reduction of uromodulin fragments (36). Additional consistent and prominent changes include the reduction of fragments of CD99 and the polymeric immunoglobulin receptor (37). Such changes have also been observed in connection with severe/lethal courses of COVID-19 and have been interpreted as possible indications of endothelial damage (30,34–37).

There are numerous efforts to predict acute kidney injury in both medically ill and severely injured patients(38). Many functional tests aim to make predictions not through the quantification of laboratory chemical parameters, but through the performance of urine output after stress tests, such as the furosemide stress test or renal functional reserve(38). The standardized furosemide stress test has a good predictive probability (AUC 0.87, p = 0.001) in a limited test group, but the testing requires a coordinated timing of furosemide administration and documentation of time and urine output(39). This must be accommodated within the time-intensive care of critically ill patients(40).

Genetic variations as determinants for both risk and outcome are not well defined(41). Association studies have identified a large number of genetic polymorphisms that are capable of predicting different and variable kidney responses to the same type of injury(41). However, identifying these individual genetic susceptibilities for AKI plays no role in managing acute courses in previously healthy, severely injured patients, or in prognostic assessment and therapy timing. Overall, the results of such models are variable and often inconsistent. The lack of robust and reproducible associations with AKI is not surprising given the complex, multifactorial nature of chronic and acute renal insufficiency(38). None of these studies combined the prognostic information from potential genetic polymorphisms with existing prediction models(38). Ideal models for such clinical studies continue to be applied in elective cardiac surgery(42). The reason for this is that cardiac surgical procedures not only represent a large surgical population but also have a well-studied epidemiology of AKI in this context, due to the use of heart-lung machines and the resultant potential ischemia- reperfusion injury to the kidney(42). However, this population cannot in any way reflect patients who suffer not only massive musculoskeletal damage but also direct injuries to the kidney

Severely injured patients with preclinical shock due to hemorrhage and extensive extremity trauma with muscle damage often suffer from kidney damage, which frequently leads to acute renal failure requiring dialysis (43). The need for a predictive tool is substantial, as the prediction and estimation of interventions such as dialysis in the daily routine of intensive care are not currently predicted by injury or mortality scores (ISS, TRISS, RISC II) or by progression assessments such as the APACHE II score. These outcomes still depend on the rapid detection of the condition and the continuous, rigorous monitoring of patients (44).

In our patient cohort, 4 patients (29%) showed acute kidney injury. This pilot study showed a significant prediction probability for the development of acute kidney injury in polytraumatized patients [CKD273 (p = 0.004) and AKI_204 (p = 0.029)].

### ARDS

Regarding ARDS and estimating interventions such as prone positioning or ECMO, we face a similar peptide deregulations to AKI (Supplementary Table 3). Papurica et al. and Cao et al. demonstrated a wide range of ARDS-relevant genes and factors (45,46). Cao et al. developed a promising prediction model that could improve early clinical management and intervention in the development of ARDS (46). However, one of the main problems in analyzing genes and microRNAs as predictive tools is the lack of standardization of methods for extraction, quantification, and data analysis (46). In contrast, the handling of urinary samples does not present major challenges, as it is an excretory product without special demands in terms of purity and concentration, making it easily obtainable (22,33,37).

In clinical practice, the Berlin Definition is currently used as the gold standard for ARDS treatment (47). It relies on various criteria that must be fulfilled to make a diagnosis (47). The severity of ARDS is determined based on the ratio of arterial oxygen partial pressure to fractional inspired oxygen (PaO2/FiO2), with different levels established depending on the measured values (47). The appearance of these criteria and the rise in the required oxygen partial pressure can sometimes occur days later (47). Often, clinicians are confronted with the clinical picture of ARDS unexpectedly. The silent period before onset can lead to an irreversible spiral, and interventions may be decided too late as a response (47).

ARDS is characterized by severe hypoxemia due to non-cardiogenic pulmonary edema (48). Both COVID-19 and ARDS frequently exhibit diffuse alveolar damage, which is marked by the destruction of the alveolar-capillary barrier (48). The presence of non-cardiogenic pulmonary edema is a hallmark of ARDS and is also observed in severe COVID-19 cases (49–51). Both conditions involve a hyperactive inflammatory response (48). In COVID-19, this is often referred to as a "cytokine storm," where excessive inflammatory cytokines cause extensive lung damage, similar to what is seen in ARDS(48). Although there are similar expressions in both diseases, the pathophysiology of COVID-19 differs from ARDS due to its diverse complexity(48,49).

Some severe COVID-19 cases develop classical ARDS, but a considerable portion of severe cases do not conform to the classical ARDS characteristics of "reduced lung volume and decreased compliance"(48–52). In these patients, pulmonary compliance is nearly normal, which is not consistent with the severity of hypoxemia(53). Furthermore, ARDS-related hypoxemia is mainly caused by intrapulmonary shunts, supplemented by dead space ventilation(54). COVID-19-related hypoxemia may be explained by a dysfunction of hypoxic pulmonary vasoconstriction, leading to a loss of lung perfusion regulation and pulmonary microthrombi(53,55).

Although the courses of Covid-19 and classical ARDS differ in some pathophysiological aspects, they do have similarities. Especially through the development of ARDS in severe Covid cases, these adverse outcomes can be identified by the urinary proteomic COV50 marker. In our study, out of 10 (71,4%) patients with thoracic trauma, 4 developed respiratory insufficiency (28,6%), of which only one patient manifested ARDS (7,1). The COV50 showed only a trend in predicting these endpoint (p = 0.094). To better assess the suitability of this urinary proteome, a larger number of patients need to be included compared to this pilot study.

Due to the small number of patients and low manifestation of ARDS at the time of the pilot study, no reliable statements could be made for prognostic assessment, treatment interventions, and intervention timing, such as determining when ECMO (7,1%) or prone positioning (7,1%) is temporally indicated. Nevertheless, considering the trend of COV50 alongside other proteomic results, it appears to be a promising avenue to address these exact issues in the management of severe ARDS cases.

### Scores

The ISS is calculated by squaring the three highest values of the AIS and then adding them together (56). It is used to quantify and assess the overall injury severity of a patient based on injuries in different body regions (56). However, the ability to determine injury severity based on the discriminative abilities of an ISS threshold depends on a reproducible definition of "severe" (57). The ISS is highly dependent on the person collecting the data (58).

Experienced individuals have recorded significantly higher values (58). This inter-observer variability often makes the score’s interpretation challenging and less explicit. Evaluating the severity of an injury (i.e., direct acute impact, excluding effects of comorbidities, complications, and treatments) depends on the specific aspects of the illness being considered (57). This evaluation can involve various parameters, such as the likelihood of death, the complexity and extent of required treatments and resources, the probability and degree of permanent impairment, and the impact on the patient’s quality of life (57).

Accordingly, in this study, we see no significant prediction of an endpoint by the ISS (p = 0.3). The TRISS optimizes the prediction probability and, as a combination score of the RTS and ISS, provides a probability of survival (59). Although historical literature shows alignment with expected predictions, our study found no significance in predicting any endpoint (p = 0.2). There is also criticism of the score, as the variables may not capture the full complexity of the patient’s condition (60).

Another approach to improving the prediction of survival probability in polytraumatized patients is the RISC II, which includes not only anatomical injury descriptions but also (patho)physiological indicators (26,57). By incorporating these discriminators, the score aims to estimate the risk of death in severely injured patients more effectively (26). Integrating the RISC II into daily intensive care practice, especially as a tool for monitoring the course of the disease, is significantly challenging due to its difficult accessibility and determination, as the score is primarily designed for initial survival probability assessment (26). Table 1 also shows no significant prediction probability for the endpoints with the RISC II (p = 0,4).

Scores specifically designed and expanded for the intensive care unit promise better predictive accuracy for critical courses. The APACHE II score, in addition to parameters from individual organ systems, incorporates age, current clinical findings, and medical history to provide a scoring system for assessing critically ill patients, allowing daily predictions about survival probability (1,17,60). Neither on the first day nor throughout the entire sampling period did the APACHE II score show a significant predictive accuracy for any endpoint in this study (p = 0.4) (Table 1, Figure 3).

The SAPS II provides an assessment of the risk of death and is a tool for predicting the intensive care trajectory of critically ill patients (13). The primary admission diagnosis does not need to be entere (13). The severity of multiple injuries does not influence the predictive accuracy (13). This is confirmed in our research question. The SAPS II score shows no significance in predicting the endpoint (p = 0.7). These results are in line to other retrospective multicenter studies, stating the APACHE II score and the SAPS II score did not show acceptable performance in predicting outcomes for intensive care trauma patients (60). Only the APACHE III score was reliable and is still frequently used in daily intensive care medicine (61). However, the score requires additional software support that compares the entered data with historical benchmark data from U.S. hospitals (61).

In our pilot study, none of the established scores showed a significant association with the combined endpoint. This is likely due to the limited power of the study. However, two of the urinary peptide classifiers, AKI204 and CKD273, significantly predicted the outcome (p = 0.029, p = 0.004).

Nevertheless, these scores are a gold standard in their respective areas of use and are employed daily in the assessment and care of severely injured and critically ill intensive care patients. In addition to predicting acute kidney injury, CKD273 significantly correlates with the ISS and its associated assessment of the severity of patients’ injuries (p = 0.034). Likewise, CKD273 shows a correlation with the APACHE II-Score, which was the only score collected at all sampling points and provides an estimate of the progression (p < 0.001). AKI_207 also showed a significant correlation with scores for the assessment of intensive care patients and their progression. The classifier correlated with the SAPS II (p = 0.031) and the APACHE II score (p < 0.001) and could provide an outlook on the intensive care course and care requirements. Similar to CKD273, Cov50 also showed a significant correlation with the ISS (p = 0.042), as well as with the RISC II (p = 0.040) and the APACHE II-Score (p < 0.001). Thus, Cov50 could also serve as a classifier for the primary assessment of injury severity and the resulting mortality risk.

### Limitations

This study has several limitations. First, the sample size was small, with a total of 16 patients and only 14 with polytrauma; thus, this study was designed as a pilot study. Additionally, the complete set of samples (14 days, 5 samples per patient) could only be collected from 8 patients. Furthermore, there were some heterogeneous samples. Due to the small number of patients, the study population exhibited a highly heterogeneous pattern of injuries and injury severity. Furthermore, not all relevant laboratory values could be collected on all sampling days. Additionally, there are challenges in data collection once patients are transferred from the ICU to the general ward.

## Conclusion

The prediction of severe organ damage/organ failure and their critical courses in severely injured patients is still associated with high uncertainty due to individual patterns of injury and progression.The prediction of these events is frequently based on the expertise and experience of the intensivist, with frequent laboratory tests and close monitoring of important organ systems, such as the liver, kidneys, heart, and lungs, required to prepare for unexpected courses and indications for interventions. Nonetheless, intensive care physicians are often faced with unforeseen developments.

This pilot study should serve as a proof of concept for applying proteome analysis in traumatology, paving the way for a multicenter study, as a significantly greater number of cases is required for a conclusive evaluation of the investigated biomarkers.

## Data Availability

The datasets generated and analysed during the current study are not publicly available but are available from the corresponding author upon reasonable request.

## List of abbreviations

AIS: Abbreviated injury scale
AKI: acute kidney injury
APACHE II: acute physiology and chronic health evaluation II
ARDS: acute respiratory distress syndrome
CE-MS: Capillary electrophoresis–mass spectrometry
CKD: Chronic kidney disease
CRP: C-Reaktive Protein
ECMO: extracorporeal membrane oxygenation
FDR: false discovery rate
GCS: Glasgow coma scale
Hb: Haemoglobin
ICU: intensive care unit
ISS: injury severity score
MODS: multiple organ dysfunction syndrome
MOD: multiorgan dysfunction
MOF: multiorgan failure
SOFA: sequential organ failure assessment
NT-proBNP: N-terminal prohormone of brain natriuretic peptide
PRBCs: packed red blood cells
PTT: partial thromboplastin time
RISC: revised injury severity classification
RTS: revised trauma score
SAPS II: simplified acute physiology score
TISS: therapeutic intervention scoring system
TRISS: trauma and injury severity score

---

**Supplementary Table 1:**
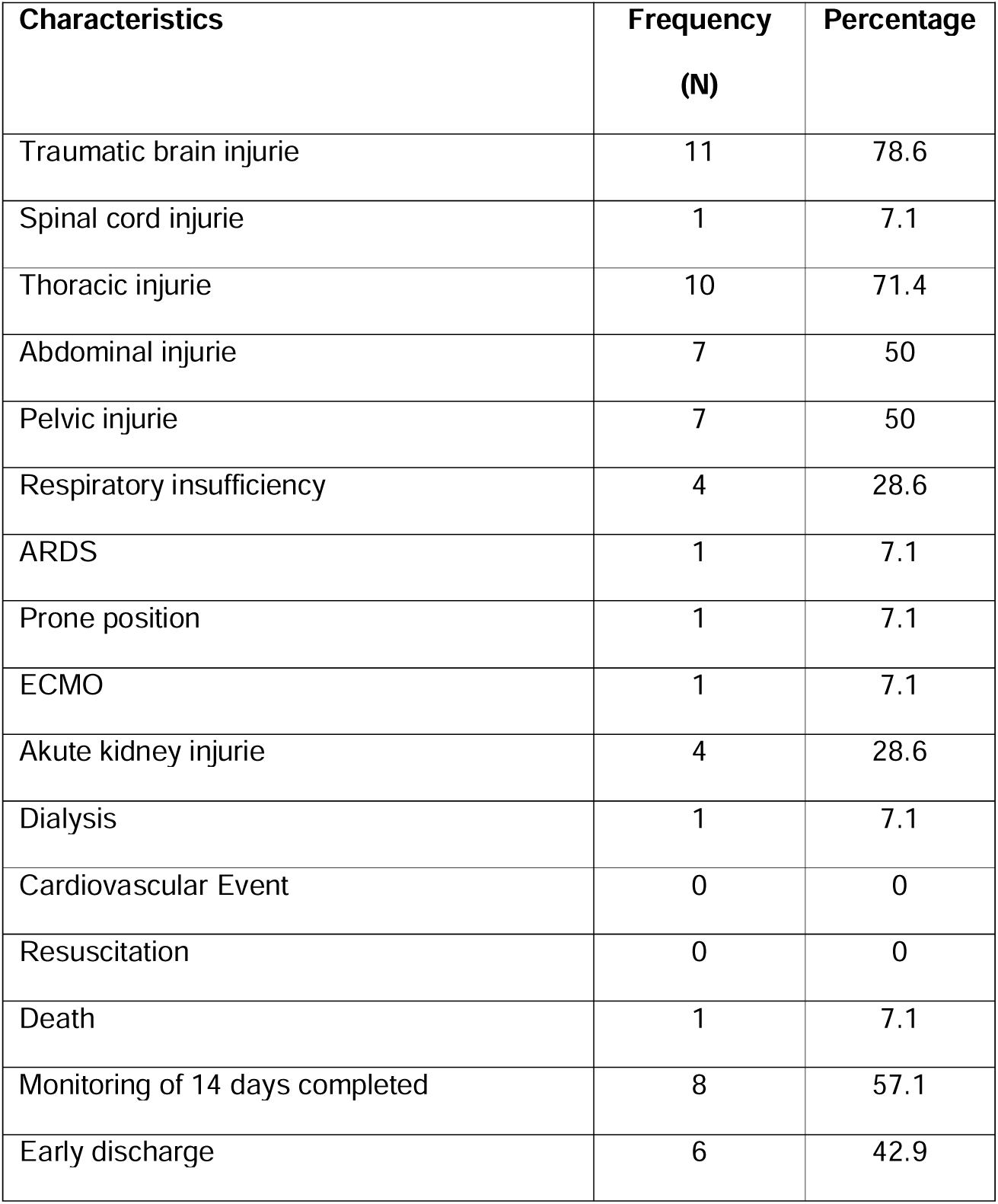
Frequencies of injuries and events in the 14 polytrauma patients in the cohort during the 14-day monitoring period.

**Supplementary Table 2:** Frequencies of successful sample collection, score and blood sample trends during the monitoring period of 14 days.

| Characteristics | Total | Frequency | Percentage |
| --- | --- | --- | --- |
| 1. Sample collection on admission | 14 | 14 | 100 |
| 2. Sample collection on day 2 | 14 | 14 | 100 |
| 3. Sample collection on day 5 | 14 | 12 | 85,7 |
| 4. Sample collection on day 10 | 14 | 9 | 64,3 |
| 5. Sample collection on day 14 | 14 | 8 | 57,1 |
| Characteristics | Total | Mean | SD |
| 1. APACHE II score on admission | 14 | 12,29 | 7,64 |
| 2. APACHE II score on day 2 | 14 | 11,21 | 6,02 |
| 3. APACHE II score on day 5 | 12 | 10,92 | 6,24 |
| 4. APACHE II score on day 10 | 9 | 12,11 | 6,21 |
| 5. APACHE II score on day 14 | 8 | 12,57 | 7,66 |
| 1. Lactate on admission | 14 | 2,76 | 1,91 |
| 2. Lactate on day 2 | 13 | 1,59 | 1,59 |
| 3. Lactate on day 5 | 10 | 0,96 | 0,35 |
| 4. Lactate on day 10 | 5 | 0,82 | 0,27 |
| 5. Lactate on day 14 | 3 | 0,73 | 0,12 |
| 1. CRP on admission | 14 | 3,20 | 5,02 |
| 2. CRP on day 2 | 14 | 169,48 | 99,46 |
| 3. CRP on day 5 | 13 | 104,01 | 90,20 |
| 4. CRP on day 10 | 9 | 138,98 | 166,64 |
| 5. CRP on day 14 | 7 | 56,39 | 38,97 |
| 1. Hb on admission | 14 | 11,69 | 2,66 |
| 2. Hb on day 2 | 14 | 9,06 | 2,18 |
| 3. Hb on day 5 | 13 | 9,02 | 1,71 |
| 4. Hb on day 10 | 9 | 8,32 | 1,32 |
| 5. Hb on day 14 | 7 | 8,89 | 0,88 |

**Supplementary Table 3:** Urinary peptides significantly different in abundance between healthy controls and trauma patients (N=16 each) (.xls)

